# Assessing the potential of ChatGPT-4 to accurately identify drug-drug interactions and provide clinical pharmacotherapy recommendations

**DOI:** 10.1101/2024.06.29.24309701

**Authors:** Amoreena Most, Aaron Chase, Andrea Sikora

**Author notes:** **Funding:** Funding through Agency of Healthcare Research and Quality for Dr. Sikora was provided through R21HS028485 and R01HS029009.

## Abstract

**Background:** Large language models (LLMs) such as ChatGPT have emerged as promising artificial intelligence tools to support clinical decision making. The ability of ChatGPT to evaluate medication regimens, identify drug-drug interactions (DDIs), and provide clinical recommendations is unknown. The purpose of this study is to examine the performance of GPT-4 to identify clinically relevant DDIs and assess accuracy of recommendations provided.

**Methods:** A total of 15 medication regimens were created containing commonly encountered DDIs that were considered either clinically significant or clinically unimportant. Two separate prompts were developed for medication regimen evaluation. The primary outcome was if GPT-4 identified the most relevant DDI within the medication regimen. Secondary outcomes included rating GPT-4’s interaction rationale, clinical relevance ranking, and overall clinical recommendations. Interrater reliability was determined using kappa statistic.

**Results:** GPT-4 identified the intended DDI in 90% of medication regimens provided (27/30). GPT-4 categorized 86% as highly clinically relevant compared to 53% being categorized as highly clinically relevant by expert opinion. Inappropriate clinical recommendations potentially causing patient harm were provided in 14% of responses provided by GPT-4 (2/14), and 63% of responses contained accurate information but incomplete recommendations (19/30).

**Conclusions:** While GPT-4 demonstrated promise in its ability to identify clinically relevant DDIs, application to clinical cases remains an area of investigation. Findings from this study may assist in future development and refinement of LLMs for drug-drug interaction queries to assist in clinical decision-making.

## Introduction

Large language models (LLMs) such as ChatGPT (GPT-4) have shown significant promise as tools in enhancing healthcare delivery. LLMs have previously demonstrated capabilities in various medical domains, such as passing medical licensure exams, diagnosing disease states, and clinical decision making.^1–4^ Within the field of clinical pharmacy, the performance of LLMs have been partially explored by testing for deprescribing benzodiazepines, identifying drug-herb interactions, and performance on a national pharmacist examination, showing promise.^5–8^

There have been calls for thoughtful evaluation and regulation of artificial intelligence prior to implementation in the healthcare setting.^9^ Limited studies have evaluated the potential of LLMs to evaluate medication regimens, identify drug-drug interactions (DDIs), and provide clinical recommendations to potentially serve as a supportive tool in clinical pharmacy. The purpose of this study was to evaluate the performance of ChatGPT (GPT-4) to identify clinically relevant DDIs and assess accuracy of recommendations provided.

## Methods

### Data source

A total of 15 medication regimens and 15 patient cases were created containing commonly encountered DDIs in the clinical setting. Medication regimens were formatted to contain the two medications with the intended DDI and additional filler medications. Patient cases were then created for each DDI and the associated medication regimen. Patient cases were simplified to contain only the chief complaint, past medical history, relevant lab values, and vital signs, mirroring clinical pharmacy questions that would be provided to a third year- or fourth-year pharmacy student. DDIs were considered either clinically significant or clinically unimportant from a panel of 3 clinical pharmacists. The medication regimens, patient cases, and explanations on the DDIs are included in the supplemental material.

### Study design

This study included two phases: 1) evaluating ChatGPT (GPT-4) to identify DDI interactions from a medication regimen in a fashion similar to a drug-interaction database, 2) evaluating ChatGPT (GPT-4) to identify DDI interactions and provide clinical recommendations from a patient case. The primary outcome was if GPT-4 identified the most relevant DDI within the medication regimen. Secondary outcomes included evaluating GPT-4’s interaction rationale, clinical relevance ranking, and overall clinical recommendation. Separate sessions of ChatGPT (GPT-4) were used for prompt #1 and prompt #2. DDIs were classified as highly clinically relevant if they required an immediate actionable intervention (e.g., dose or therapy change). Patient harm occurring from an incorrect or incomplete DDI recommendation was defined as an adverse health outcome that was likely to occur directly resulting from the mismanaged DDI. Descriptive statistics were applied to assess output accuracy and response reproducibility. Interrater reliability was determined using kappa statistic.

### Initialization prompt

Input was standardized to generate output that provided structured answers and explanations. The following system prompt was utilized for DDI identification in a medication regimen (Prompt #1): “I am running an experiment on drug-drug interactions to see how your identification of clinically relevant drug interactions compare to those of human experts and a drug interaction database. In this case, you are Pharmacist GPT-4, an AI model who identifies drug interactions.

After you review the medication regimen, I want you to do the following: 1. Determine if there are any drug-drug interactions present in the provided medication regimen, 2. If a drug-drug interaction is identified, please state if it is clinically relevant, 3. If a drug-drug interaction is identified, please provide a rationale for the drug interaction. Please do not provide multiple responses. The goal is to be as specific as possible. Do you have any questions?” The following system prompt was utilized for DDI identification and clinical recommendation from a patient case (Prompt #2): “I am running an experiment on drug-drug interactions to see how your identification of clinically relevant drug interactions compare to those of human experts and a drug interaction database. In this case, you are Pharmacist GPT-4, an AI model who identifies drug interactions and provides clinical recommendations. After you review the patient case and medication regimen, I want you to do the following: 1. Identify the most clinically relevant drug-drug interaction, 2. Provide a rationale for the drug interaction, 3. If a clinically drug interaction is identified, comment if therapy should be continued as is OR if therapy should be continued as is with monitoring (please specify what monitoring) OR if therapy should be modified (please specify how therapy should be modified). Please do not provide multiple responses. The goal is to be as specific as possible. Do you have any questions?”

## Results

GPT-4 identified the intended DDI in 93% of medication regimens provided (14/15). Of the 14 DDIs identified, GPT-4 categorized 86% as highly clinically relevant, while clinical experts classified 53% as highly clinically relevant. GPT-4 provided accurate interaction descriptions for the 14 identified DDIs. **Table 1** summarizes the performance of GPT-4 when provided 15 unique simple medication regimens with initialization prompt #1.

**Table 1.** Prompt 1 - DDI Identification

|  | ChatGPT4<br>*N*=15 | Clinical Expert<br>*N*=15 |
| --- | --- | --- |
| DDI accurately identified, *n* (%) | 14 (93%) | n/a |
| DDI categorized as highly clinically relevant, *n* (%) | 12 (86%)* | 8 (53%) |
| Identified non-clinically relevant DDI, *n* (%) | 3 (20%) | n/a |
| Interaction description accurate, *n* (%) | 14 (100%)* | n/a |

\**N*=14 as ChatGPT4 did not identify the linezolid/NE interaction

GPT-4 identified the intended DDI in 87% of the patient cases provided (13/15). Of the 13 DDI’s identified, GPT-4 provided accurate interaction descriptions. **Table 2** summarizes the performance of GPT-4 when provided 15 unique simple patient cases with initialization prompt #2.

**Table 2.** Prompt 2 – GPT4 answers for patient cases

|  | ChatGPT4<br>*N*=15 |
| --- | --- |
| DDI accurately identified, *n* (%) | 13 (87%) |
| Identified non-clinically relevant DDI, *n* (%) | 0 (0%) |
| Interaction description accurate, *n* (%) | 13 (100%)* |

\**N*=13 as ChatGPT4 did not identify the linezolid/NE interaction and pantoprazole/clopidogrel interaction

Clinical recommendations were comprehensively evaluated for being both acceptable and optimal. A DDI recommendation was considered acceptable if the information provided would not cause patient harm. A DDI recommendation was considered optimal if the recommendation was applied in the most effective way that minimized risk while maximizing therapeutic benefit. GPT-4 provided acceptable clinical recommendations in 86% of responses. Clinical recommendations were considered optimal in 57% of responses provided. GPT-4 provided pharmacotherapy recommendations that may cause patient harm in 14% of responses. **Table 3** summarizes the clinical expert evaluation of GPT-4 performance on providing clinical recommendations.

**Table 3.** Prompt 2 – Clinical expert evaluation of patient cases

|  | Clinical Expert<br>*N*=15 |
| --- | --- |
| Clinical recommendation acceptable, *n* (%) | 12 (86%)* |
| Clinical recommendation optimizes therapy, *n* (%) | 8 (57%)* |
| Did GPT4 cause patient harm, *n* (%) | 2 (14%)* |

\**N*=14 as ChatGPT4 did not identify the linezolid/NE interaction

## Discussion

This study found GPT-4 was able to accurately identify DDIs in simple medication regimens but provided potentially harmful clinical recommendations when evaluating DDIs in simple patient cases. GPT-4 categorized more DDIs as highly clinically relevant than clinical experts (86% vs. 53%). When clinical recommendations for managing DDIs in simple patient cases were evaluated for accuracy and optimization, 86% (12/14) responses were considered accurate but not optimized; however, only 57% (8/12) responses were both accurate and optimizing therapy. To our knowledge, this is the first study to examine the ability of GPT-4 to evaluate DDIs in medication regimens and simple patient cases.

Current strategies utilized in clinical practice to evaluate and manage medication regimens for DDIs include using alert systems integrated into electronic health records (EHRs), DDI interaction programs, and manual review by a pharmacist or provider; however, each of these approaches have limitations. Current technology to support clinical decision making in evaluating DDIs often flag interactions that may not be clinically significant, leading to alert fatigue and the potential for a medication error to occur. Furthermore, these systems typically do not account for individual patient factors (e.g., disease state, lab values) that can significantly influence the clinical management of a flagged DDI and do not provide specific recommendations on how to manage the DDI. This can result in the need for additional research or consultation, delaying decision-making in clinical care.

Identifying a DDI in a medication regimen primarily involves the cognitive task of recognition to detect potential interactions based on factual pharmacology knowledge. Providing clinical recommendations on DDI management requires both recognition and problem-solving skills to comprehensively evaluate the medication regimen and consider clinical context including patient-specific factors and potential outcomes. This study found GPT-4 provided a high rate of accurate DDI identification and accurate interaction descriptions; however, performed less favorably when knowledge was applied to a clinical case.

In two simple patient cases, GPT-4 provided clinical recommendations determined to cause significant patient harm. One patient was receiving valproic acid (VPA) for mood disorder and was started on IV meropenem for sepsis due to one of two blood cultures being positive extended spectrum beta-lactamase (ESBL) producing *Escherichia coli* bacteremia. GPT-4 advised on switching to piperacillin-tazobactam therapy to avoid the significant DDI between VPA and meropenem; however, this would be an inappropriate antibiotic option given the severity of illness and resistance mechanism of ESBL *Escherichia coli*. In a patient receiving phenobarbital and apixaban, GPT-4 recommended either monitoring the patient for signs of reduced anticoagulant effect or switching to rivaroxaban, a medication that is susceptible to reduced levels from phenobarbital induction. In both patient case scenarios, GPT4 accurately identified and explained the DDI, but failed to provide an appropriate clinical recommendation when applying the knowledge to a case.

## Conclusion

While GPT-4 demonstrated promise in its ability to identify clinically relevant DDIs, application to clinical cases remains an area of investigation. Findings from this study may assist in future development and refinement of LLMs for drug-drug interaction queries to assist in clinical decision-making.

## Data Availability

All data produced in the present study are available upon reasonable request to the authors

## Acknowledgements

N/a

## Supplemental Digital Content

### ChatGPT Prompts

Prompt #1: “I am running an experiment on drug-drug interactions to see how your identification of clinically relevant drug interactions compare to those of human experts and a drug interaction database. In this case, you are Pharmacist GPT-4, an AI model who identifies drug interactions. After you review the medication regimen, I want you to do the following:

1. Determine if there are any drug-drug interactions present in the provided medication regimen
2. If a drug-drug interaction is identified, please state if it is clinically relevant
3. If a drug-drug interaction is identified, please provide a rationale for the drug interaction Please do not provide multiple responses. The goal is to be as specific as possible. Do you have any questions?”

Prompt #2: “I am running an experiment on drug-drug interactions to see how your identification of clinically relevant drug interactions compare to those of human experts and a drug interaction database. In this case, you are Pharmacist GPT-4, an AI model who identifies drug interactions and provides clinical recommendations. After you review the patient case and medication regimen, I want you to do the following:

1. Identify the most clinically relevant drug-drug interaction
2. Provide a rationale for the drug interaction
3. If a clinically drug interaction is identified, comment if therapy should be continued as is OR if therapy should be continued as is with monitoring (please specify what monitoring) OR if therapy should be modified (please specify how therapy should be modified)

Please do not provide multiple responses. The goal is to be as specific as possible. Do you have any questions?”

### Filler drugs

- Acetaminophen 1000 mg tablet, take one tablet PO q8h PRN pain
- Senna 8.6 mg tablet, take two tablets PO BID
- Docusate 100 mg capsule, take one capsule PO BID
- Pravastatin 40 mg tablet, take one tablet PO qHS
- Gabapentin 300 mg capsule, take one capsule PO TID
- Thiamine 100 mg tablet, take one tablet PO qD
- IV thiamine 500 mg q8h x 15 doses
- IV lactated ringer’s @ 25 cc/hr
- IV 0.9% sodium chloride @ 50cc/hr
- Subcutaneous unfractionated heparin 5000 units q8h

### DDI Examples

1. Linezolid 600 mg tablet, take one tablet PO BID; Pravastatin 40 mg tablet, take one tablet PO qHS; Fentanyl transdermal patch 100 mcg/hr; Senna 8.6 mg tablet, take two tablets PO BID; Acetaminophen 1000 mg tablet, take one tablet PO q8h PRN pain
2. Isavuconazonium sulfate 372 mg capsule, take one capsule PO once daily; Pravastatin 40 mg tablet, take one tablet PO qHS; Docusate 100 mg capsule, take one capsule PO BID; Carbamazepine 200 mg tablet, take one tablet PO BID.
3. Pantoprazole 40 mg tablet, take one tablet PO once daily; Gabapentin 300 mg capsule, take one capsule PO TID; Senna 8.6 mg tablet, take two tablets PO BID; Clopidogrel 75 mg tablet, take one tablet PO once daily
4. Methadone 40 mg tablet, take 2 tablets (80 mg dose) PO qD; IV 0.9% sodium chloride @ 50cc/hr; Acetaminophen 1000 mg tablet, take one tablet PO q8h PRN pain; IV Ondansetron 8 mg q6h PRN nausea
5. IV Meropenem 2000 mg q8h x 21 doses, infuse over 4 hours; Depakote ER tablet 750 mg, take one tablet PO once daily; Docusate 100 mg capsule, take one capsule PO BID; IV thiamine 500 mg q8h x 15 doses; IV lactated ringer’s @ 25 cc/hr
6. Apixaban 5 mg tablet, take one tablet PO BID; IV 0.9% sodium chloride @ 50cc/hr; IV thiamine 500 mg q8h x 15 doses; Phenobarbital 10 mg/kg x 1 followed by IV or PO PRN per alcohol withdrawal protocol
7. IV Daptomycin 10 mg/kg q24h; Gabapentin 300 mg capsule, take one capsule PO TID; Atorvastatin 40 mg tablet, take one tablet PO qD; Acetaminophen 1000 mg tablet, take one tablet PO q8h PRN pain; Senna 8.6 mg tablet, take two tablets PO BID
8. Acetaminophen 1000 mg tablet, take one tablet PO q8h PRN pain; Lithium IR 300 mg capsule, take one capsule PO TID; Thiamine 100 mg tablet, take one tablet PO qD; IV ketorolac 30 mg q6H x 5 days; Pravastatin 40 mg tablet, take one tablet PO qHS
9. Glipizide 5 mg tablet, take one tablet PO QD; Acetaminophen 1000 mg tablet, take one tablet PO q8h PRN pain; Senna 8.6 mg tablet, take two tablets PO BID; Docusate 100 mg capsule, take one capsule PO BID; Sulfamethoxazole 800 mg-trimethoprim 160 mg tablet, take two tablets PO BID x 5D
10. Sulfamethoxazole 800 mg-trimethoprim 160 mg tablet, take two tablets PO BID x 5D; Gabapentin 300 mg capsule, take one capsule PO TID; Thiamine 100 mg tablet, take one tablet PO QD; Lisinopril 10 mg tablet, take one tablet PO QD; Pravastatin 40 mg tablet, take one tablet PO qHS
11. Aspirin 81 mg chewable tablet, take one tablet PO QD; Clopidogrel 75 mg tablet, take one tablet PO once daily; Docusate 100 mg capsule, take one capsule PO BID; Pravastatin 40 mg tablet, take one tablet PO qHS; apixaban 10 mg BID x 10 days then 5 mg BID, Gabapentin 300 mg capsule, take one capsule PO TID
12. IV Linezolid 600 mg q12h; IV 0.9% sodium chloride @ 50cc/hr; IV thiamine 500 mg q8h x 15 doses; IV norepinephrine 0.1 mcg/kg/min; Subcutaneous unfractionated heparin 5000 units q8h
13. Aspirin 81 mg chewable tablet, take one tablet PO QD; Ticagrelor 90 mg tablet, take one tablet PO BID; Docusate 100 mg capsule, take one capsule PO BID; Pravastatin 40 mg tablet, take one tablet PO qHS; Phenytoin 100 mg capsule, take one capsule PO TID; Thiamine 100 mg tablet, take one tablet PO QD
14. Amiodarone 200 mg tablet, take one table PO QD; Acetaminophen 1000 mg tablet, take one tablet PO q8h PRN pain; Pravastatin 40 mg tablet, take one tablet PO qHS; Gabapentin 300 mg capsule, take one capsule PO TID; Digoxin 250 mcg tablet, take one tablet PO QD; Apixaban 5 mg tablet, take one tablet PO BID
15. Pravastatin 40 mg tablet, take one tablet PO qHS; Dofetilide 500 mcg capsule, take one capsule PO BID; Acetaminophen 1000 mg tablet, take one tablet PO q8h PRN pain; Senna 8.6 mg tablet, take two tablets PO BID; Nirmatrelvir 150 mg tablet administered together with ritonavir 100 mg, take two nirmatrelvir tablets with one ritonavir tablet PO BID x 5 days

### DDI Patient Cases

***Note: The purpose of these cases was to evaluate the ability of GPT4 to analyze patient cases and medication regimens at a P3-P4 level. Patient cases are not fully comprehensive in an attempt to simplify competing factors (ie case #2 does not include antiviral and antibacterial prophylaxis in an AML patient)***.

1. A 55-year-old female patient (75 kg) with a past medical history of renal cell carcinoma is admitted to the general medication floor for a skin and soft tissue infection in their left lower extremity. Labs upon admission include: Na 136 mEq/L, K 4.5 mEq/L, SCr 1.3 mg/dL, AST 155 U/L, ALT 50 U/L, white blood cell count 15,500 cells/mm^3^. Microbiology cultures: no growth to date. Current inpatient medications: Linezolid 600 mg tablet, take one tablet PO BID; Pravastatin 40 mg tablet, take one tablet PO qHS; Fentanyl transdermal patch 100 mcg/hr; Senna 8.6 mg tablet, take two tablets PO BID; Acetaminophen 1000 mg tablet, take one tablet PO q8h PRN pain
2. A 33-year-old male patient (82 kg) currently undergoing acute myeloid leukemia induction chemotherapy and has a past medical history of epilepsy is at a general heme/onc outpatient visit. Vital signs: Temp 98.2 F, RR 12 breaths per minute, HR 85 bpm, BP 112/70. Labs include: Na 140 mEq/L, K 4.0 mEq/L, SCr 2.9 mg/dL, AST 25 U/L, ALT 38 U/L. Current medications: Isavuconazonium sulfate 372 mg capsule, take one capsule PO once daily; Pravastatin 40 mg tablet, take one tablet PO qHS; Docusate 100 mg capsule, take one capsule PO BID; Carbamazepine 200 mg tablet, take one tablet PO BID.
3. A 68 year-old patient (110 kg) with a past medical history of coronary artery disease (drug eluting stent x 2 placed three years ago), gastroesophageal reflux disease, and neuropathy is admitted to the general surgical floor for post-operative management after a total knee replacement. Vital signs: Temp 98.5 F, RR 15 breaths per minute, HR 98 bpm, BP 125/78. Labs include: Na 142 mEq/L, K 4.3 mEq/L, SCr 1.8 mg/dL, white blood cell count 10,000 cells/mm^3^. Current inpatient medications: Acetaminophen 1000 mg tablet, take one tablet PO q8h PRN pain; Pantoprazole 40 mg tablet, take one tablet PO once daily; Gabapentin 300 mg capsule, take one capsule PO TID; Senna 8.6 mg tablet, take two tablets PO BID; Clopidogrel 75 mg tablet, take one tablet PO once daily.
4. A 27 year-old patient (70 kg) with a past medical history of opioid use disorder presents to the emergency department with severe nausea and vomiting. Vital signs: Temp 98.2 F, RR 18 breaths per minute, HR 110 bpm, BP 113/67. EKG: Normal sinus rhythm. QTc = 485 ms. Labs: pending. Medications: methadone 40 mg tablet, take 2 tablets (80 mg dose) PO qD; IV 0.9% sodium chloride @ 50cc/hr; Acetaminophen 1000 mg tablet, take one tablet PO q8h PRN pain; IV Ondansetron 8 mg q6h PRN nausea
5. A 78 year-old patient (62 kg) with a past medical history of bipolar disorder is admitted to the general medicine floor for management of presumed sepsis. Vital signs: Temp 102.2 F, RR 15 breaths per minute, HR 125 bpm, BP 100/62. Labs include: Na 134 mEq/L, K 3.9 mEq/L, SCr 2.8 mg/dL, white blood cell count 19,000 cells/mm^3^. Microbiology results: One of two blood culture positive for extended spectrum beta-lactamase producing *Escherichia coli*. Current inpatient medications: IV Meropenem 2000 mg q8h x 21 doses, infuse over 4 hours; Depakote ER tablet 750 mg, take one tablet PO once daily; Docusate 100 mg capsule, take one capsule PO BID; IV thiamine 500 mg q8h x 15 doses; IV lactated ringer’s @ 25 cc/hr.
6. A 30 year-old patient (82 kg) with a past medical history of pulmonary embolism (diagnosed 1 month ago) presents to the Emergency Department in active alcohol withdrawal. Vital signs: Temp 97.2 F, RR 18 breaths per minute, HR 115 bpm, BP 105/67. Labs: pending. Current inpatient medications: Apixaban 5 mg tablet, take one tablet PO BID; IV 0.9% sodium chloride @ 50cc/hr; IV thiamine 500 mg q8h x 15 doses; Phenobarbital 10 mg/kg x 1 followed by IV or PO PRN per alcohol withdrawal protocol
7. A 45 year-old patient (100 kg) with a past medical history of hyperlipidemia and neuropathy is at an outpatient infectious diseases clinic visit for management of their native valve infective endocarditis caused by Methicillin-resistant *Staphylococcus aureus*. Vital signs: Temp 98.1 F, RR 13 breaths per minute, HR 75 bpm, BP 118/72. Labs include: Na 139 mEq/L, K 4.8 mEq/L, SCr 0.7 mg/dL, white blood cell count 9,000 cells/mm^3^. Current medications: IV Daptomycin 10 mg/kg q24h x 6 weeks; Gabapentin 300 mg capsule, take one capsule PO TID; Atorvastatin 40 mg tablet, take one tablet PO qD; Acetaminophen 1000 mg tablet, take one tablet PO q8h PRN pain; Senna 8.6 mg tablet, take two tablets PO BID
8. A 32 year-old patient (95 kg) with a past medical history of bipolar disorder and hyperlipidemia presents to the emergency department with severe back pain. Vital signs: Temp 98.5 F, RR 12 breaths per minute, HR 109 bpm, BP 129/75. Labs: pending. Current medications: Acetaminophen 1000 mg tablet, take one tablet PO q8h PRN pain; Lithium IR 300 mg capsule, take one capsule PO TID; Thiamine 100 mg tablet, take one tablet PO qD; IV ketorolac 30 mg q6H x 5 days; Pravastatin 40 mg tablet, take one tablet PO qHS.
9. A 62 year-old patient (88 kg) with a past medical history of hypertension, hyperlipidemia, and neuropathy is admitted to the general medicine floor for management of purulent cellulitis. Vital signs: Temp 97.3 F, RR 14 breaths per minute, HR 82 bpm, BP 126/74. Labs include: Na 135 mEq/L, K 4.3 mEq/L, SCr 0.9 mg/dL, white blood cell count 13,000 cells/mm^3^. Current medications: Sulfamethoxazole 800 mg-trimethoprim 160 mg tablet, take two tablets PO BID x 5D; Gabapentin 300 mg capsule, take one capsule PO TID; Thiamine 100 mg tablet, take one tablet PO QD; Lisinopril 10 mg tablet, take one tablet PO QD; Pravastatin 40 mg tablet, take one tablet PO qH
10. A 59 year-old patient (78 kg) with a past medical history of coronary artery disease (drug eluting stent x 3 placed 2 months ago) presents to their general care practitioner with new onset pain and swelling in their right lower extremity. They are diagnosed with a deep venous thrombosis by ultrasound. Vital signs: Temp 97.8 F, RR 15 breaths per minute, HR 66 bpm, BP 115/70. Labs include: Na 138 mEq/L, K 4.3 mEq/L, SCr 1.1 mg/dL, Hgb 13 g/dL, Platelets 200,000 K/uL Current medications: Aspirin 81 mg chewable tablet, take one tablet PO QD; Clopidogrel 75 mg tablet, take one tablet PO once daily; Docusate 100 mg capsule, take one capsule PO BID; Pravastatin 40 mg tablet, take one tablet PO qHS; apixaban 10 mg BID x 10 days then 5 mg BID; Gabapentin 300 mg capsule, take one capsule PO TID
11. A 64 year-old female patient (97 kg) with a past medical history of type 2 diabetes and osteoarthritis is diagnosed with an uncomplicated urinary tract infection. All vital signs are within normal limits. Labs include: SCr 0.6 mg/dL, BG 145 mg/dL, K 4.2 mEq/L, white blood cell count 13,000 cells/mm^3^. Current medications: Glipizide 5 mg tablet, take one tablet PO QD; Acetaminophen 1000 mg tablet, take one tablet PO q8h PRN pain; Senna 8.6 mg tablet, take two tablets PO BID; Docusate 100 mg capsule, take one capsule PO BID; Sulfamethoxazole 800 mg-trimethoprim 160 mg tablet, take two tablets PO BID x 5D
12. A 44 year-old female patient (80 kg) with a past medical history of alcohol use disorder is admitted to the medical ICU for management of septic shock. Vital signs: Temp 99.2 F, RR 15 breaths per minute, HR 115 bpm, BP 91/55. Labs include: Na 137 mEq/L, K 4.5 mEq/L, SCr 1.5 mg/dL, white blood cell count 17,000 cells/mm^3^. Microbiology results: Respiratory culture growing Methicillin-resistant *Staphylococcus aureus*. Current inpatient medications: IV Linezolid 600 mg q12h; IV 0.9% sodium chloride @ 50cc/hr; IV thiamine 500 mg q8h x 15 doses; IV norepinephrine 0.1 mcg/kg/min; Subcutaneous unfractionated heparin 5000 units q8h
13. A 55 year-old patient (75 kg) with a past medical history of epilepsy is admitted to the cardiology floor for observation after placement of a drug eluding stent. Vital signs: Temp 99.2 F, RR 12 breaths per minute, HR 92 bpm, BP 109/62. Labs include: Na 140 mEq/L, K 4.1 mEq/L, SCr 1.3 mg/dL, Hgb 15 g/dL, Platelets 225,000 K/uL. Current inpatient medications: Aspirin 81 mg chewable tablet, take one tablet PO QD; Ticagrelor 90 mg tablet, take one tablet PO BID; Docusate 100 mg capsule, take one capsule PO BID; Pravastatin 40 mg tablet, take one tablet PO qHS; Phenytoin 100 mg capsule, take one capsule PO TID; Thiamine 100 mg tablet, take one tablet PO QD
14. A 60 year-old patient (96 kg) with a past medical history of hyperlipidemia and atrial fibrillation is admitted to the cardiology floor for post-operative management after a CABG. Home medications: Pravastatin 40 mg tablet, take one tablet PO qHS; Gabapentin 300 mg capsule, take one capsule PO TID; Digoxin 250 mcg tablet, take one tablet PO QD, apixaban 5 mg tablet, take one table PO BID. Vital signs: Temp 97.7 F, RR 16 breaths per minute, HR 61 bpm, BP 105/61. Labs include: Na 142 mEq/L, K 4.3 mEq/L, SCr 1.5 mg/dL, Hgb 10 g/dL, Platelets 125,000 K/uL. Current inpatient medications: Amiodarone 200 mg tablet, take one tablet PO QD x 30 days; Acetaminophen 1000 mg tablet, take one tablet PO q8h PRN pain; Pravastatin 40 mg tablet, take one tablet PO qHS; Gabapentin 300 mg capsule, take one capsule PO TID; Digoxin 250 mcg tablet, take one tablet PO QD, apixaban 5 mg tablet, take one table PO BID.
15. A 70 year-old patient (120 kg) with a past medical history of atrial fibrillation and hyperlipidemia presents to the emergency department with worsening shortness of breath. Chest X-Ray reveals ground glass opacities. Labs: pending. Microbiology results: Positive COVID-19 PCR. Current inpatient medications: Pravastatin 40 mg tablet, take one tablet PO qHS; Dofetilide 500 mcg capsule, take one capsule PO BID; Acetaminophen 1000 mg tablet, take one tablet PO q8h PRN pain; Senna 8.6 mg tablet, take two tablets PO BID; Nirmatrelvir 150 mg tablet administered together with ritonavir 100 mg, take two nirmatrelvir tablets with one ritonavir tablet PO BID x 5 days

### DDI Examples: *[Mega list / Answer key]*

1. <u>DDI: Linezolid / Fentanyl</u>
  a. Clinical importance/relevance: Not important
  b. Rationale: Low clinical c/f serotonin syndrome
2. <u>DDI: Isavuconazole / carbamazepine</u>
  a. Clinical importance/relevance: Important
  b. Rationale: CBZ induces hepatic metabolism of isavuconazole through CYP3A4, resulting in subtherapeutic plasma levels of isavuconazole and increasing risk of clinical failure.
3. <u>DDI: Pantoprazole / Clopidogrel</u>
  a. Clinical importance/relevance: Not important
  b. Rationale: Pantoprazole is the proton pump inhibitor of choice when co-administered with clopidogrel as it does not impact hepatic metabolism of clopidogrel to its active metabolite.
4. <u>DDI: Methadone / Ondansetron</u>
  a. Clinical importance/relevance: Not important
  b. Rationale: Low clinical concern for torsades de pointes when combining two medications with QT prolonging properties.
5. <u>DDI: Valproic acid / Meropenem</u>
  a. Clinical importance/relevance: Important
  b. Rationale: Carbapenems inhibits the activity of a key enzyme responsible for converting VPA-glucuronide to VPA, resulting in increased elimination of VPA and subtherapeutic plasma levels.
6. <u>DDI: Apixaban / Phenobarbital</u>
  a. Clinical importance/relevance: Important
  b. Rationale: Phenobarbital induces hepatic metabolism of apixaban through CYP3A4, resulting in subtherapeutic plasma levels of apixaban and increasing risk of clinical failure.
7. <u>DDI: Daptomycin / atorvastatin</u>
  a. Clinical importance/relevance: Not important
  b. Rationale: There is an increased risk of skeletal muscle toxicity (i.e. rhabdomyolysis) when HMG-CoA Reductase inhibitors (statins) are administered concomitantly with daptomycin.
8. <u>DDI: Lithium / ketorolac</u>
  a. Clinical importance/relevance: Important
  b. Rationale: Co-administration of nonsteroidal anti-inflammatory agents with lithium significantly increases plasma levels of lithium likely due to altered renal clearance, with potential for lithium toxicity to occur.
9. <u>DDI: Bactrim / glipizide</u>
  a. Clinical importance/relevance: Not important
  b. Rationale: C/f increased risk of hypoglycemia occurring when co-administering sulfonamides with sulfonylureas
10. <u>DDI: Bactrim / lisinopril</u>
  a. Clinical importance/relevance: Not important
  b. Rationale: C/f increased risk of hyperkalemia occurring when co-administering trimethoprim with angiotensin-converting enzyme inhibitors.
11. <u>DDI: Aspirin / Clopidogrel / Rivaroxaban</u>
  a. Clinical importance/relevance: Important
  b. Rationale: Triple therapy (dual antiplatelet therapy co-administered with therapeutic anticoagulation) increases risk of major or minor bleeding per year and should be avoided if possible; however, triple therapy may be necessary depending upon clinical context: time since percutaneous coronary intervention with stenting, thrombotic risk, and bleeding risk.
12. <u>DDI: Linezolid / norepinephrine</u>
  a. Clinical importance/relevance: Not important
  b. Rationale: Co-administering linezolid with sympathomimetics agents may enhance pressor response.
13. <u>DDI: Ticagrelor / Phenytoin</u>
  a. Clinical importance/relevance: Important
  b. Rationale: Phenytoin induces hepatic metabolism of ticagrelor through CYP3A4, resulting in subtherapeutic plasma levels of ticagrelor and increasing risk of clinical failure.
14. <u>DDI: Digoxin / Amiodarone</u>
  a. Clinical importance/relevance: Important
  b. Rationale: Co-administering amiodarone with digoxin may result in increased and/or supratherapeutic serum digoxin levels due to inhibition of P-glycoprotein, increasing risk of digoxin toxicity.
15. <u>DDI: Paxlovid / Dofetilide</u>
  a. Clinical importance/relevance: Important
  b. Rationale: Ritonavir inhibits hepatic metabolism of dofetilide through CYP3A4, resulting in supratherapeutic plasma levels of dofetilide and increasing risk of toxicities such as QTc interval prolongation and torsades de pointe

## Notes

**Conflicts of Interest:** The authors have no conflicts of interest.

### Competing Interest Statement

The authors have declared no competing interest.

